# Identifying single-nucleotide polymorphisms (SNPs) intersecting Alzheimer’s disease pathology and end-of-life traits using genomic informational field theory (GIFT)

**DOI:** 10.64898/2026.03.05.26347710

**Authors:** Sam Heysmond, Panagiota Kyratzi, Jonathan Wattis, Andras Paldi, Keeley J Brookes, Karim Kreft, Beili Shao, Cyril Rauch

## Abstract

Quantitative genome-wide association studies (GWAS) typically use additive linear models to assess average phenotypic differences between genotype groups. While effective for identifying common variants in large cohorts, these methods compress detailed phenotypic data into summary statistics, potentially limiting detection of subtle genotype–phenotype relationships. Genomic Informational Field Theory (GIFT) is a newly developed framework that preserves fine-grained trait structure by analysing ranked phenotypic configurations rather than relying on group means. We applied GIFT to genetic and neuropathological data from the Brains for Dementia Research cohort and directly compared its performance with conventional GWAS. Principal component analysis (PCA) was used to derive independent quantitative traits linked to Alzheimer’s disease neuropathology (CERAD, Thal phase, and Braak staging), with and without inclusion of age at death. Both GWAS and GIFT were conducted on the same filtered genotype dataset. Both approaches identified genome-wide significant associations (p<10⁻⁶) within the APOE locus, consistent with established Alzheimer’s disease risk factors. However, GIFT detected additional significant SNPs not identified by GWAS, implicating genes involved in amyloid processing, neuronal apoptosis, synaptic function, neuroinflammation, and metabolic regulation. GIFT also identified loci associated with variation in age at death that were not detected by GWAS, highlighting pathways linked to lipophagy, mitochondrial quality control, sphingolipid metabolism, frailty, and ageing. These findings demonstrate that rethinking analytical representation, rather than solely increasing sample size, can substantially expand the discovery potential of genetic association studies, offering a transparent and complementary framework for quantitative genomics in deeply phenotyped datasets.

**Authors summary:** The genetic basis of complex traits and diseases is highly polygenic, and robust genotype–phenotype associations typically require large sample sizes. Genomic Informational Field Theory (GIFT) is a recently developed analytical approach that combines non-parametric, rank-based statistics with pattern-based analysis to infer genotype–phenotype relationships. By preserving the relative structure and distribution of genomic signals at SNP-level, GIFT has the potential to detect associations in small cohorts. Initially developed in the field of animal genetics, where studies often involve limited sample sizes, we apply GIFT here for the first time in human genetics, specifically to the Brains for Dementia Research cohort, and compare its performance with conventional genome-wide association studies (GWAS). We conclude that GIFT constitutes a complementary and interpretable methodology capable of enhancing inference in contexts where large sample sizes are impractical but phenotypic resolution is high, thereby helping to mitigate longstanding power limitations in quantitative genomics.

## Introduction

Traditional Genome-Wide Association Studies (GWAS) assume an additive inheritance model with a binary trait or diagnosis in a logistic regression model to compare the average risk expressed as an odds ratio between genotypes of each single nucleotide polymorphism (SNP). Analysis for quantitative traits uses the sample principle with a linear regression, comparing the averages of the measured trait between genotype groups, but also provides increased statistical power to detect SNP associations for the same underlying genetic effect, due to the greater information about the variability between individuals. Therefore, utilising measures of underlying pathology of diseases might uncover more of the genetic aetiology. However, the use of averages in GWAS to compare genotypic groups inherently restricts information on individual variance and is primarily designed to detect additive linear genetic effects. Given the small effect sizes typically associated with genetic variants influencing complex traits or diseases, GWAS may fail to capture subtler effects.

Recently, a new methodology for identifying SNPs associated with traits has emerged. Known as Genomic Informational Field Theory (GIFT) and inspired by field theory in physics and information theory, this approach preserves the informational content of datasets through an alternative representation and analysis of datasets.

Traditional statistics is underpinned by frequency- or count-based visualizations, such as bar charts or histograms, which group raw data into discrete bins. When these discrete representations are interpolated in the continuum limit and conceptualized as continuous distributions, they are referred to as distribution density functions (DDFs). A key example of DDF is the normal distribution, widely employed in GWAS. Ultimately, DDFs interpreted within the frequentist probability framework provide the foundation for calculating key population-level statistical summaries (e.g., averages, variances, and measures of significance) that subsequently underpin population-level inferences. However, while DDF-based statistics and related statistical summaries reveal population-level properties, they do so at a cost: by transforming raw data into aggregated representations, the process sacrifices information about the highly resolved structure and fine-grained diversity of individual datapoints. To illustrate this point, consider a population in which phenotypic measurements are captured with high precision, rendering each data point unique, a scenario increasingly frequent with the advent of advanced measurement technologies (1). Once these data are grouped into bins to construct frequency- or count-based plots, the individuality of each measurement is effectively erased, as all points within a bin become indistinguishable. In effect, data binning results in a substantial loss of the information originally present in the dataset’s unaggregated form. Consequently, although the statistical summaries derived from DDFs may appear informative, they are ultimately products of information loss rooted in a categorized representation of the original dataset, which imposes limits on the knowledge that can be extracted from the data. Therefore, GIFT treats each quantitative trait measurement as a unique data point and analyses it according to its rank order within the dataset, preserving individual-level information and enabling more nuanced insights. GIFT then determines the association between genotype and phenotype by computing the cumulative sum of genotypes corresponding to the ranked trait values, from small to large trait values. The cumulative sum of genotypes reported to the position in the rank defines a curve termed ‘genetic path’, and noted *θ* thereafter, captures not only the effects linked to genotypes averages (allele substitution) but also the effects linked their variances (2–5). The significance of the association is then determined by comparing the maximal amplitude difference of the observed genetic path, *θ*, with that of the resulting average null-path, noted *θ̄*_0_, generated using the same method but with the ranked trait values and corresponding genotypes randomly permuted an infinite number of times. Formalised theoretically (3–5), GIFT’s ability to detect genotype–phenotype associations has been evaluated using simulations (3), and real-world datasets (2). These studies demonstrate its capacity to detect small effect sizes without requiring sample sizes in the thousands.

The aim of this study was to investigate the effectiveness of PCA analysis from ordinal scale pathology categories and co-variates to increase the range of unique datapoints and therefore information to be used in GWAS and GIFT, and subsequently compare the genetic associations identified from both analyses. This investigation utilised genetic data from the Brains for Dementia Research (BDR) cohort (6), including detailed neuropathological assessments and age at death (n = 563). The study compared genetic association findings derived from conventional GWAS and GIFT across two related datasets. The first dataset comprised variables specifically reflecting Alzheimer’s disease (AD) neuropathology, while the second incorporated both AD neuropathological measures and age at death. This design aimed to determine whether genetic variants associated with AD pathology also contribute to variation in lifespan, or whether distinct genetic factors influence survival independently of AD-related pathological processes. We demonstrate that only GIFT identified genes *(NECTIN2*, *TOMM40*, and *APOE*) were jointly associated with both AD neuropathology and age at death. In contrast, a substantial number of additional loci were associated with either AD pathology or age at death independently, but not with both traits simultaneously.

## Results

### Principles underscoring GIFT in genotype-phenotype associations

GIFT is an optimal methodology for identifying associations between datasets without relying on predefined categories or data grouping. To illustrate its approach, consider a continuously varying trait measured with sufficient precision such that no two phenotypic values are identical. In biallelic organisms, three genotypic categories, bb, bB/Bb, and BB, are present, following Mendelian inheritance. Assuming the trait is linked to these genotypes, potential associations can be explored using count or frequency plots, as shown in Figs 1A and 1B. These plots typically reveal that each genotype occurs predominantly within specific ranges of the continuous trait, a pattern that can be confirmed using statistical tests comparing the mean trait values, as shown in Fig 1C.

**Figure 1.**
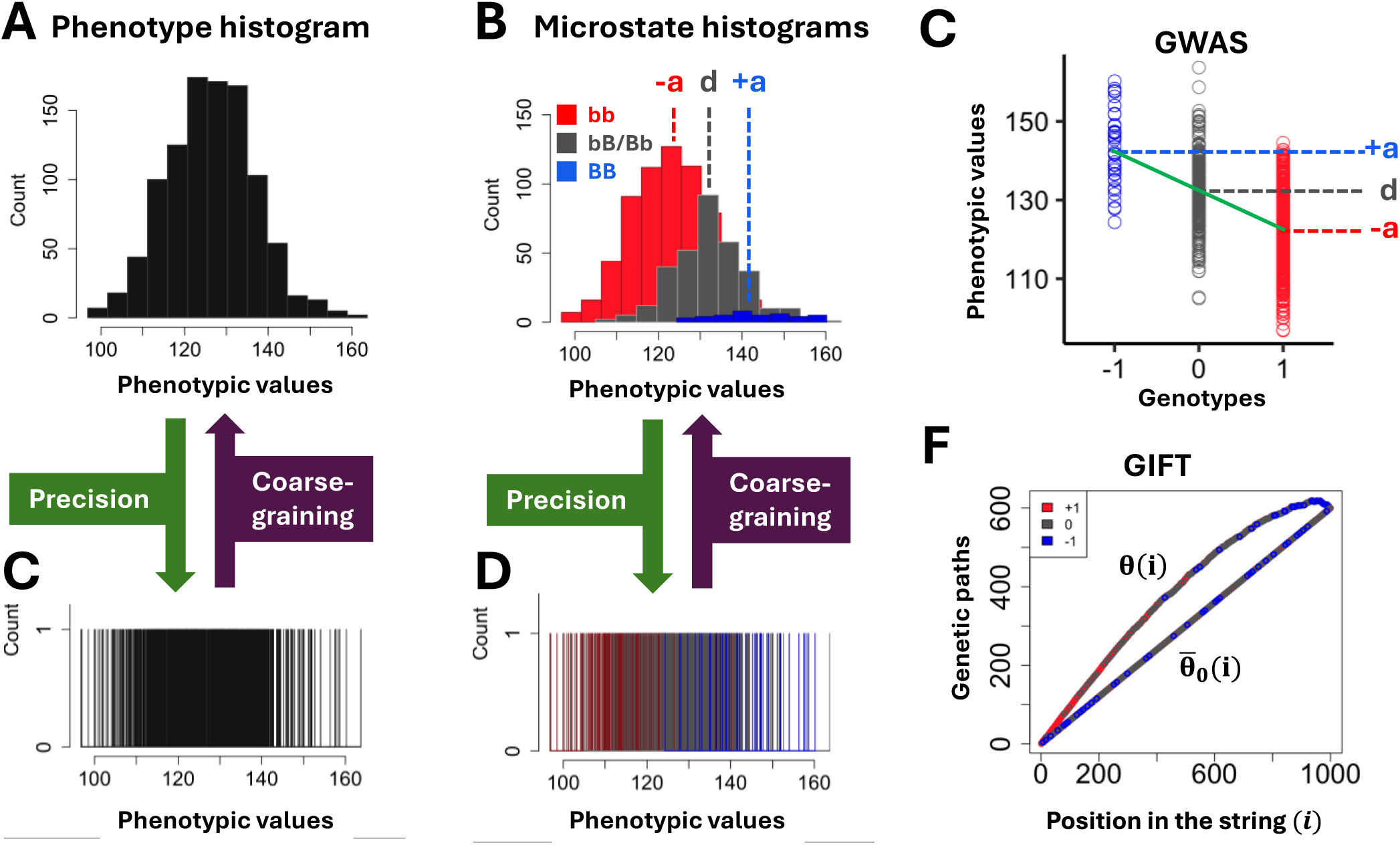
GWAS and GIFT representations of associations: GWAS and GIFT simulation using 1,000 individuals, with a gene effect (2a) equal to the standard deviation of the phenotype distribution, zero dominance, and genotype frequencies consistent with Hardy–Weinberg equilibrium (HWE). **(A)** Representation of the phenotype distribution using a histogram (count plot). **(B)** Representation of genotype frequencies using count plots. In biallelic (b, B) organisms, three genotypes are observed: bb, bB/Bb, and BB. **(C)** In the GWAS framework, genotype–phenotype associations are evaluated by comparing the mean phenotypic values associated with each genotype (−a, d, +a), allowing estimation of the statistical significance of the association, the gene effect size (2a), and dominance (d). **(D–E)** As measurement precision increases, fewer data points fall within each bin, and at full precision the histogram effectively resolves into barcode-like representations. Crucially, coarse-graining GIFT’ s detailed outputs (barcodes) recover GWAS representational framework **(F)** GIFT determines genotype–phenotype associations by evaluating the probability of specific genotype configurations, introducing the concept of genetic paths to infer statistical significance.

Nevertheless, this approach may mask subtle associations that GIFT is specifically designed to reveal. One major drawback of categorising continuous data is the loss of detailed information, as differences within each category cannot be captured. GIFT addresses this issue by examining associations without grouping the data, preserving its full resolution and allowing for more accurate inferences, even with smaller sample sizes.

As shown in Figs 1D and 1E, if we were to know the exact value of datapoints, the absence of categories compels us to concentrate on barcodes to provide means to extract inferences. Importantly, coarse graining the barcodes recover the traditional way of representing datasets under the form of histograms. Now, concentrating on the different colours of the bars in Fig 1E, what is visible is that the distribution of colours is not uniform but that bars are segregated as a function of their colour, reflecting the positioning of the different distributions in Fig 1B. Importantly, while each bar is associated with a specific trait value in Fig 1E, this specific trait value depends on the subpopulation used and a different subpopulation would have provided a slightly different trait value for each bar. That is to say that what matters to determine inferences when considering the coloured barcode in Fig 1E is not the exact positioning of bars, but their relative positioning.

To determine whether the segregation of coloured bars has any significance, GIFT proceeds initially by numbering the different colours using +1 for red bars (genotype bb), 0 for gray bars (genotypes bB/Bb) and –1 for blue bars (genotype BB), thereby creating an initial string of numbers only containing +1, 0 and –1 as they appear from low to high trait values. In a second step, GIFT creates another string where the component, say at position i, corresponds to the cumulative sum of numbers located between the positions 1 to i from the initial string. The component of this new string for the position i is noted θ(i) and call genetic path. Then θ(i) is plotted as a function of the position i as shown in Fig 1F. The curved aspect of θ(i) results the presence of red bars (+1 bars) initially followed by grey bars (0 bars) and finally blue bars (-1 bars) as shown in Fig 1E.

This curve is then compared to another function resulting from a scenario corresponding to the null hypothesis. This scenario is obtained by randomly permutating the number +1, 0 and –1 in the initial string. We shall call this randomly permutated string a null string noted θ_0_(i). However, as there are many random permutations possible of the initial string one may determine the resulting average null string after considering all possible permutations. In this context, it is possible to demonstrate that the resulting average null string leads, after considering the average cumulative sum of components, to a straight-line noted θ̄ (i) (see Fig 1F), whose equation is given by, 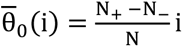, where N_+_, N_ are the number of +1, -1 bars, and N is total number of bars in the initial string, respectively. Note that the horizontal bar on the top of θ_0_ signifies that the average has been considered.

The notion of significance with GIFT is then determined by comparing the difference, θ(i) − θ̄ (i) (Fig 1F). There are different ways to determine a measure of significance for the difference θ(i) − θ̄ (i), which ultimately depend on theoretic considerations such as symmetries and invariances of the dataset and scientific problem studied. However, one generic way to determine a measure of significance is to recall that the segregation of microstates leads to large amplitudes for θ(i). One may then concentrate on the difference between the max and min values of, θ(i) − θ̄ (i) and determine the level of significance using general probability distribution theory (4).

As will be demonstrated below in the context of AD, GIFT can identify associations as long as the genotypes are not randomly distributed; in other words, unlike traditional mapping methods (GWAS) only focusing on genotypic average differences, GIFT extracts significant associations that are invisible to GWAS.

### Demographics of the BDR cohort

A subset of 563 samples of the BDR cohort was used in this study. Overall, the average age at death was 84.1 years (SD 8.9) and with 48.3% of the cohort being female. One hundred and seventy-four of these samples were defined as controls with no cognitive deficit and no dementia relevant pathology detected (mean age at death = 84.7 (SD=10.0); 52.9% female). Whilst the remaining n=389 samples displayed cognitive deficits and had AD neuropathology confirmed at post-mortem (mean age at death = 83.8 (SD=8.3); 46.3% female), no significant differences (p>0.05) were observed for these characteristics between the two groups. European ancestry of the BDR sample was confirmed previously by Chaudhury *et al.* (2019) with principal component analysis, as all samples clustered accordingly with European samples from the 1000G dataset (15).

### Principal Component Analyses

Principal Component Analysis (PCA) is a statistical dimensionality-reduction technique that transforms correlated variables into a smaller set of uncorrelated variables called principal components. Although the number of components initially equals the number of original variables, they are ordered according to the amount of variance (information) they capture from the data. Consequently, only the components that explain the largest proportion of variance are typically retained for analysis. Figs 2A and 2B show the fraction of variance explained when using AD neuropathology features (see Materials and Methods) alone and when combining AD neuropathology features with age at death, respectively. In Fig 2A, the first principal component (PC1) explains 84.6% of the variance, indicating that it is the most informative component. However, in Fig 2B, both PC1 and PC2 must be considered, as they explain 64.1% and 24.9% of the variance, respectively, together accounting for 89% of the total variance explained, thereby capturing a level of information similar to that of PC1 in Fig 2A.

**Figure 2.**
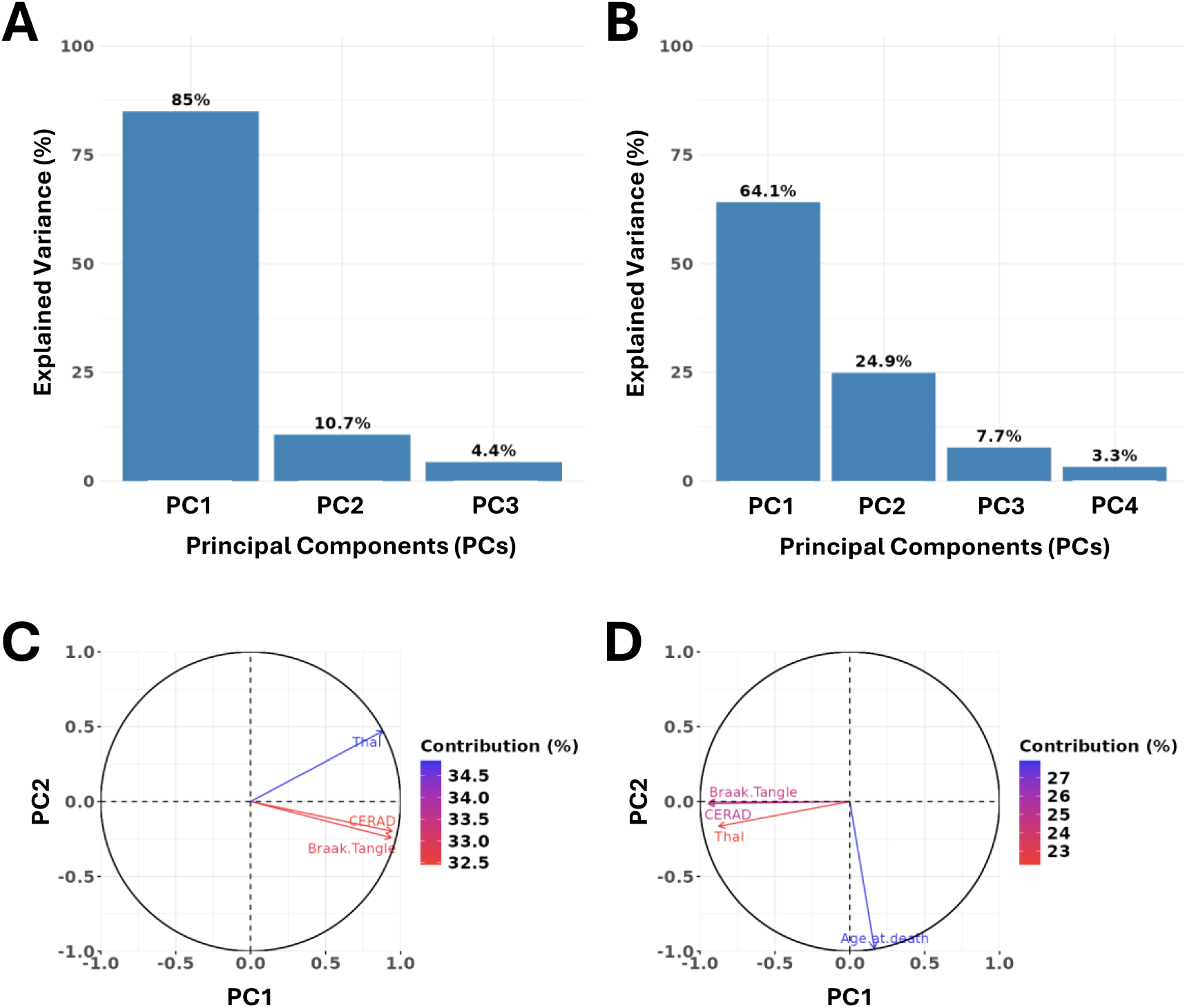
Principal components analyses. **(A)** Fraction of variances explained by the different components using the AD neuropathology variables dataset. **(B)** Fraction of variances explained by the different components using the AD neuropathology and age at death variables. **(C)** Representation of AD neuropathology vectors along the principal components PC1 and PC2. **(D)** Representation of AD neuropathology and age at death vectors along the principal components PC1 and PC2. Note: coloured labels on each bar in (C) and (D) show rounded calculations of variance explained by the components.

To determine how the initial variables distribute across the principal components, biplot representations were used for each dataset. In a biplot, the original variables are depicted as vectors emanating from the origin, where each vector corresponds to one variable. The direction of each vector indicates the variable’s influence on the principal components, while its length reflects the strength of this contribution. The angle between vectors conveys the correlation among variables: acute angles suggest positive correlation, obtuse angles indicate negative correlation, and orthogonal vectors imply no correlation. In this representation, Fig 2C shows that the AD neuropathology features are much more closely aligned with the horizontal axis (PC1) than with the vertical axis (PC2), thereby underpinning the strong correlations among AD neuropathology features, as expected from Fig 2A. In contrast, Fig 2D shows that the AD neuropathology features and age at death are aligned along different principal components, with AD neuropathology features associated with PC1 (horizontal axis) and age at death associated with PC2 (vertical axis), indicating that AD neuropathology features and age at death are, statistically, poorly correlated in this dataset.

In the remainder of the paper, we use the notation PC1AD to denote the first principal component derived from AD neuropathology variables only, and the notations PC1AD+Age and PC2AD+Age to denote the first and second principal components, respectively, derived from AD neuropathology variables together with age at death.

### GWAS and GIFT Manhattan plots

Because individual data points from the two datasets are informatively projected along their principal components, their coordinates can be used as numerical inputs to define what we term PC-related trait values. Since AD neuropathology variables inform only a single principal component (PC1AD), whereas inclusion of age at death informs two principal components (PC1AD+Age and PC2AD+Age) that are weakly correlated, two complementary analytical approaches can be pursued: one focusing on the two first principal components PC1AD and PC1AD+Age that, by virtue of Fig 2, primarily capture AD neuropathology–related variations, and another focusing on the second component PC2AD+Age, which is more strongly associated with age at death.

**#**Fig 3 shows the Manhattan plots obtained using GWAS and GIFT. Specifically, Fig 3A presents the Manhattan plot for AD neuropathology variables only (PC1AD). Fig 3B shows the Manhattan plot for the first principal component derived from AD pathology and age at death variables (PC1AD+Age), while Fig 3C displays the Manhattan plot for the second principal component (PC2AD+Age) of these same variables. Table 1 and Table 2 provide detailed information on the set of significant SNPs determined by GWAS and GIFT, respectively.

**Figure 3.**
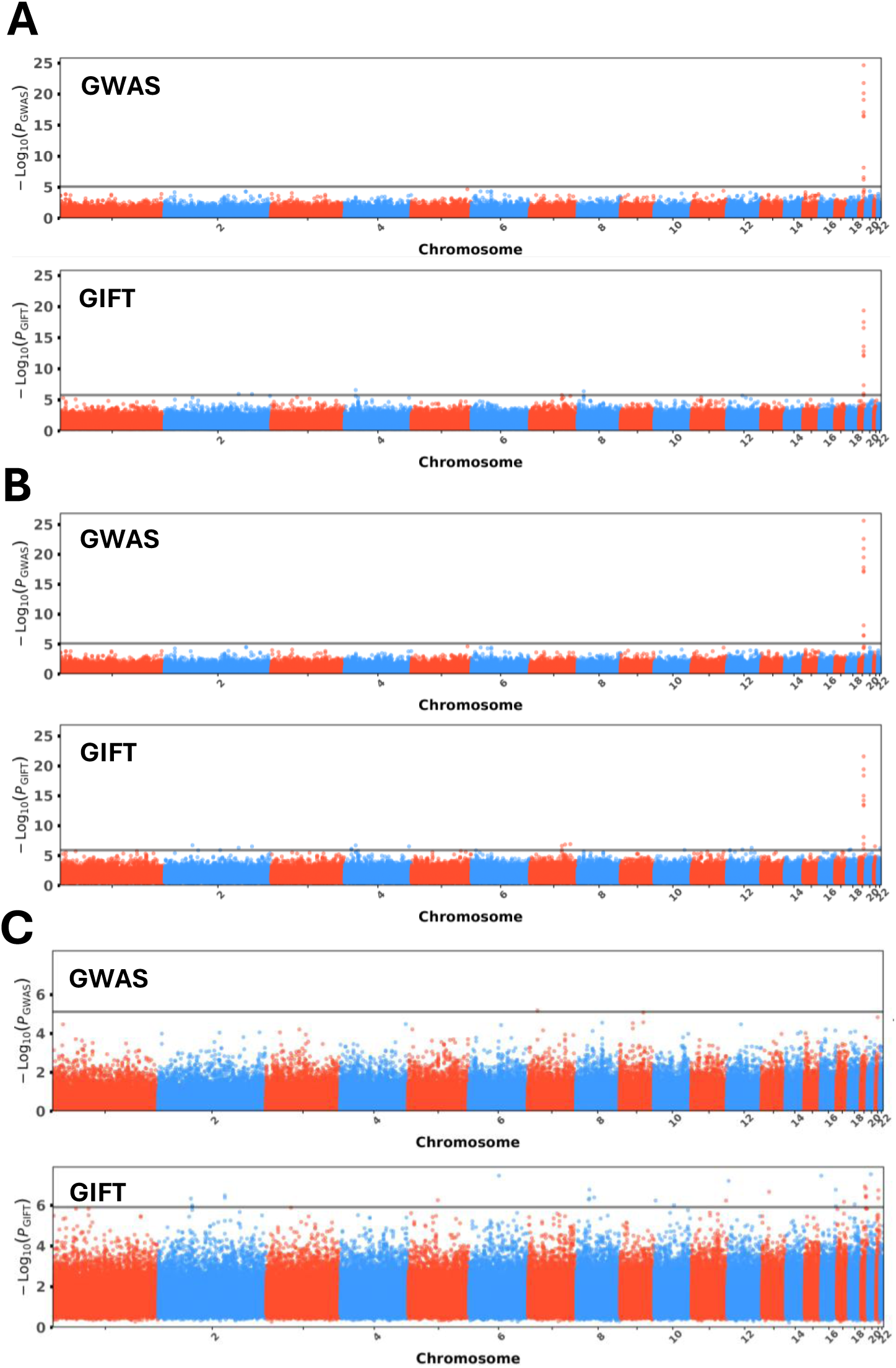
GWAS and GIFT Manhattan plots: **(A)** Genome-wide association for AD pathology variables (PC1AD). **(B)** Genome-wide association for AD pathology and age at death variables (PC1AD+Age). **(C)** Genome-wide association for AD pathology and age at death variables (PC2AD+Age). The horizontal grey line corresponds to Bonferroni correction (5%).

**Table 1:**
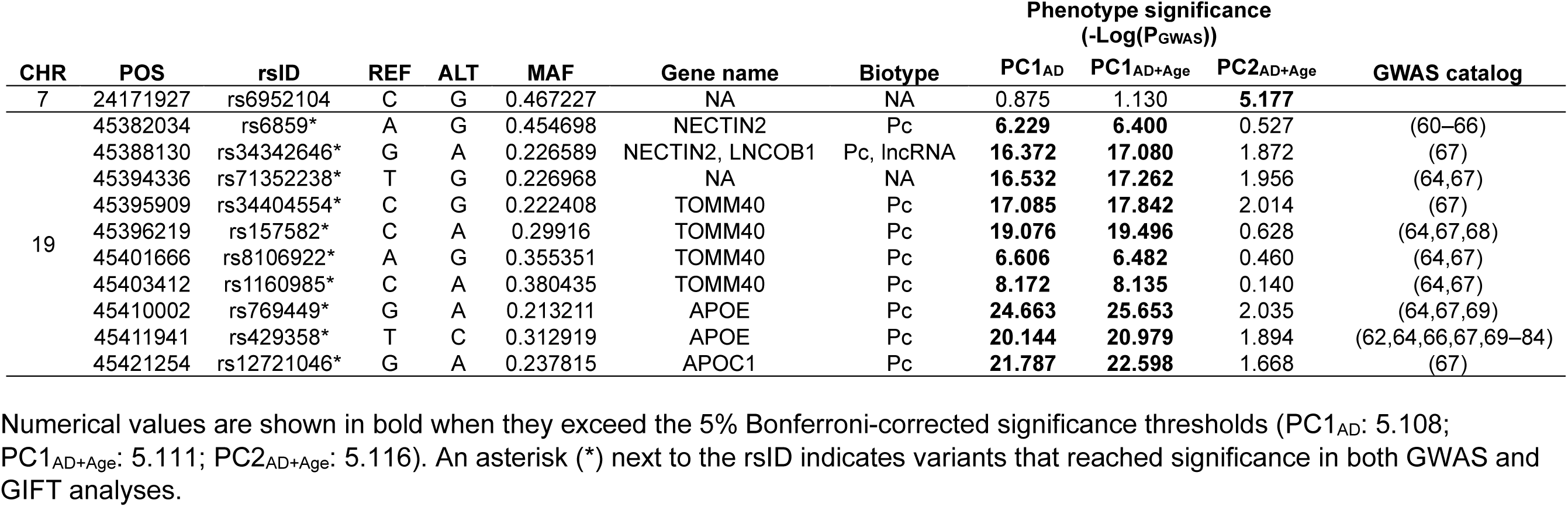
Information on significant SNPs determined by GWAS.

**Table 2:** Information on significant SNPs determined by GIFT.

| CHR | POS | rsID | REF | ALT | MAF | Gene name | Biotype | Phenotype significance: -Log(P<sub>GIFT</sub>) |  |  |
| --- | --- | --- | --- | --- | --- | --- | --- | --- | --- | --- |
|  |  |  |  |  |  |  |  | PC1<sub>AD</sub> | PC1<sub>AD+Age</sub> | PC2<sub>AD+Age</sub> |
| 2 | 51317471 | rs6734633 | G | A | 0.301003 | NRXN1-DT, NRXN1 | lncRNA, Pc | 5.344 | **6.731** | 2.053 |
|  | 59903052 | rs2110330 | A | G | 0.420435 | NA | NA | 1.206 | 1.592 | **6.329** |
|  | 63007059 | rs10153716 | A | G | 0.305695 | EHBP1 | Pc | 0.466 | 0.971 | **5.929** |
|  | 63073194 | rs11682530 | G | A | 0.307886 | EHBP1 | Pc | 0.511 | 1.146 | **5.993** |
|  | 123524253 | rs6541859 | A | G | 0.156355 | NA | NA | 4.294 | **5.912** | 1.536 |
|  | 146737198 | rs16825854 | G | A | 0.116415 | NA | NA | 0.429 | 0.960 | **6.487** |
|  | 146835932 | rs10496995 | C | A | 0.119765 | NA | NA | 0.705 | 1.208 | **6.371** |
|  | 168710172 | rs10200961 | A | G | 0.124161 | B3GALT1, B3GALT1-AS1 | Pc, lncRNA | **5.947** | **6.308** | 3.864 |
|  | 205177698 | rs7603542 | C | A | 0.11204 | NA | NA | **5.922** | **6.525** | 1.103 |
| 4 | 16281681 | rs11721706 | A | G | 0.144295 | TAPT1-AS1 | lncRNA | 3.601 | **6.136** | 1.070 |
|  | 16365531 | rs16893404 | G | A | 0.142977 | NA | NA | 3.405 | **5.960** | 0.983 |
|  | 26501114 | rs11733591 | C | A | 0.069398 | NA | NA | **6.577** | **6.706** | 1.836 |
|  | 186714443 | rs10866283 | G | A | 0.098662 | SORBS2 | Pc | 5.299 | **6.533** | 1.239 |
| 5 | 88009593 | rs254778 | G | A | 0.313758 | MEF2C-AS2 | lncRNA | 0.607 | 1.079 | **6.2523** |
| 6 | 85347257 | rs13217066 | C | A | 0.406355 | NA | NA | 2.335 | 4.057 | **7.4584** |
| 7 | 103659343 | rs17158237 | C | A | 0.109532 | NA | NA | 5.760 | **6.6287** | 0.862 |
|  | 103670208 | rs7799089 | A | G | 0.11204 | NA | NA | 5.155 | **5.9452** | 0.892 |
|  | 116435123 | rs13239139 | A | G | 0.175042 | MET | Pc | 4.446 | **6.846** | 1.361 |
|  | 136782819 | rs11761344 | C | A | 0.123746 | NA | NA | 5.609 | **6.918** | 1.481 |
| 8 | 10627844 | rs7826180 | G | A | 0.239531 | SOX7-AS1, PINX1 | lncRNA, Pc | **6.372** | 5.695 | 1.396 |
|  | 25557962 | rs11997651 | T | G | 0.275084 | NA | NA | 1.685 | 1.917 | **6.282** |
|  | 26867904 | rs2874755 | C | A | 0.0752508 | NA | NA | 0.960 | 1.480 | **6.332** |
|  | 27057879 | rs12548107 | G | A | 0.0643813 | NA | NA | 0.821 | 0.977 | **6.769** |
|  | 53445338 | rs13282005 | T | C | 0.10134 | NA | NA | 1.686 | 1.866 | **6.390** |
| 10 | 3815292 | rs2171302 | C | A | 0.16806 | NA | NA | 0.569 | 0.819 | **6.233** |
|  | 72713252 | rs827284 | T | A | 0.0991597 | SGPL1, LINC02622 | Pc, lncRNA | 3.605 | 4.331 | **6.008** |
|  | 115788094 | rs11196597 | G | A | 0.107023 | NA | NA | 4.629 | **5.947** | 0.781 |
| 11 | 131355411 | rs7951332 | G | A | 0.0543478 | NTM | Pc | 1.000 | 0.962 | **6.230** |
| 12 | 4475194 | rs10744643 | C | G | 0.383585 | NA | NA | 0.732 | 0.994 | **7.201** |
|  | 10462516 | rs2270238 | C | A | 0.328859 | KLRD1 | Pc | 4.448 | **5.918** | 1.292 |
|  | 63129713 | rs7312352 | A | G | 0.119565 | PPM1H | Pc | 5.676 | **6.023** | 1.747 |
|  | 101375823 | rs17030838 | A | G | 0.184783 | ANO4 | Pc | 3.353 | **6.304** | 1.727 |

**Table 2 (continued):** Information on significant SNPs determined by GIFT
| CHR | POS | rsID | REF | ALT | MAF | Gene name | Biotype | Phenotype significance: -Log(P<sub>GIFT</sub>) |  |  |
| --- | --- | --- | --- | --- | --- | --- | --- | --- | --- | --- |
|  |  |  |  |  |  |  |  | PC1<sub>AD</sub> | PC1<sub>AD+Age</sub> | PC2<sub>AD+Age</sub> |
| 13 | 46162672 | rs7329523 | T | G | 0.132107 | ERICH6B | Pc | 2.127 | 2.230 | **6.661** |
| 16 | 1498951 | rs12932780 | C | A | 0.101171 | CLCN7 | Pc | 2.703 | 3.208 | **7.454** |
|  | 86347358 | rs2696819 | G | A | 0.262982 | NA | NA | 1.328 | 1.694 | **6.766** |
|  | 86357163 | rs8059183 | G | A | 0.218487 | NA | NA | 0.946 | 1.138 | **5.952** |
| 17 | 50617026 | rs1492230 | A | G | 0.330268 | NA | NA | 1.487 | 2.378 | **6.167** |
| 18 | 25569058 | rs2298574 | A | G | 0.103015 | CDH2 | Pc | 3.787 | **6.027** | 1.321 |
|  | 41183021 | rs1830221 | G | A | 0.352349 | NA | NA | 1.481 | 1.852 | **6.049** |
| 19 | 34595690 | rs11881397 | G | A | 0.0854271 | NA | NA | 2.615 | 3.235 | **6.928** |
|  | 45382034 | rs6859* | A | G | 0.454698 | NECTIN2 | Pc | **5.996** | **6.947** | 2.644 |
|  | 45388130 | rs34342646* | G | A | 0.226589 | NECTIN2, LNCOB1 | Pc, lncRNA | **12.085** | **13.494** | **6.473** |
|  | 45394336 | rs71352238* | T | G | 0.226968 | NA | NA | **12.191** | **13.387** | **6.411** |
|  | 45395909 | rs34404554* | C | G | 0.222408 | TOMM40 | Pc | **12.828** | **14.233** | **6.411** |
|  | 45396219 | rs157582* | C | A | 0.29916 | TOMM40 | Pc | **13.587** | **15.015** | 4.817 |
|  | 45401666 | rs8106922* | A | G | 0.355351 | TOMM40 | Pc | 5.750 | **6.192** | 2.895 |
|  | 45403412 | rs1160985* | C | A | 0.380435 | TOMM40 | Pc | **7.347** | **8.108** | 2.723 |
|  | 45410002 | rs769449* | G | A | 0.213211 | APOE | Pc | **19.370** | **21.597** | **6.823** |
|  | 45411941 | rs429358* | T | C | 0.312919 | APOE | Pc | **17.512** | **19.446** | 4.974 |
|  | 45421254 | rs12721046* | G | A | 0.237815 | APOC1 | Pc | **16.565** | **18.396** | 5.861 |
| 20 | 17775259 | rs6080834 | G | A | 0.257525 | NA | NA | 2.173 | 2.215 | **7.526** |
| 21 | 24951271 | rs2828286 | A | G | 0.144054 | NA | NA | 4.656 | **6.547** | 0.675 |
|  | 34279458 | rs7283473 | G | A | 0.31689 | EPCIP-AS1 | lncRNA | 1.532 | 2.089 | **6.348** |
|  | 34292163 | rs7277038 | C | A | 0.317726 | EPCIP-AS1 | lncRNA | 1.326 | 1.837 | **6.739** |
| 22 | 43049014 | rs5758837 | G | A | 0.371237 | CYB5R3 | Pc | 4.329 | **6.226** | 1.369 |

Numerical values are shown in bold when they exceed the 5% Bonferroni-corrected significance thresholds (PC1<sub>AD</sub>: 5.762; PC1<sub>AD+Age</sub>: 5.909 and PC2<sub>AD+Age</sub>: 5.912). An asterisk (\*) next to the rsID indicates variants that reached significance in both GWAS and GIFT analyses.

Analysis of AD neuropathology variables only (PC1AD) identified 10 SNPs associated with the trait all within 42kb of each other around the *APOE* locus in GWAS, and at the exclusion of rs8106922 (Chr19) all of these were picked up in the GIFT analysis of the same variable. GIFT did not identify additional markers in this region but did observe significant association with an additional 4 SNPs at the 5% Bonferroni-corrected significance threshold, in *B3GALT1* (Chr2); the *SOX7-AS1/PINX1* locus (Chr8); and 2 intergenic SNPs in Chr 2 and 4 (see Tables 1 & 2).

The analysis of the neuropathology variables including age-at-death yielded PC1AD+Age and PC2AD+Age for analysis correlated with neuropathological measures and age at death, respectively.

GWAS analysis identified the same 10 SNPs as in the PC1AD analysis from the neuropathology only, with no additional SNPs, whilst GIFT identified one additional SNP, rs8103922, in the *APOE* locus to the previous pathology only results. In addition, GIFT picked up 3 of the 4 SNPs identified in the neuropathology only analysis, with rs7826180 (*SOX7-AS1/PINX*) just missing the significance cut-off. Furthermore, another 16 SNPs were found to be significantly associated with PC1AD+Age (Table 2).

Finally, analysis of PC2AD+Age derived from the AD pathological features plus age-at-death yielded only a single finding from GWAS in an intergenic region on Chr7. In contrast, GIFT identified 29 SNPs associated with this measure, highlighting multiple genes across several chromosomes (Table 2). None of these SNPs had been associated with PC1AD or PC1AD+Age measures with GIFT, expect for the 4 located in the *APOE* region (rs34342646, rs71352238, rs34404554 and rs769449), which were associated in both GIFT and GWAS and across all three PCs investigated.

### Genetic Path Information from GIFT

To clarify why SNPs identified as significant by GWAS were associated only with AD pathology features and not when age at death was included, we compared the representational frameworks of GWAS and GIFT for rs769449 (Chr19), the most significant SNP identified by GWAS and GIFT (Tables 1 & 2), as an example. Fig 4 illustrates the methodological differences between GWAS and GIFT.

**Figure 4.**
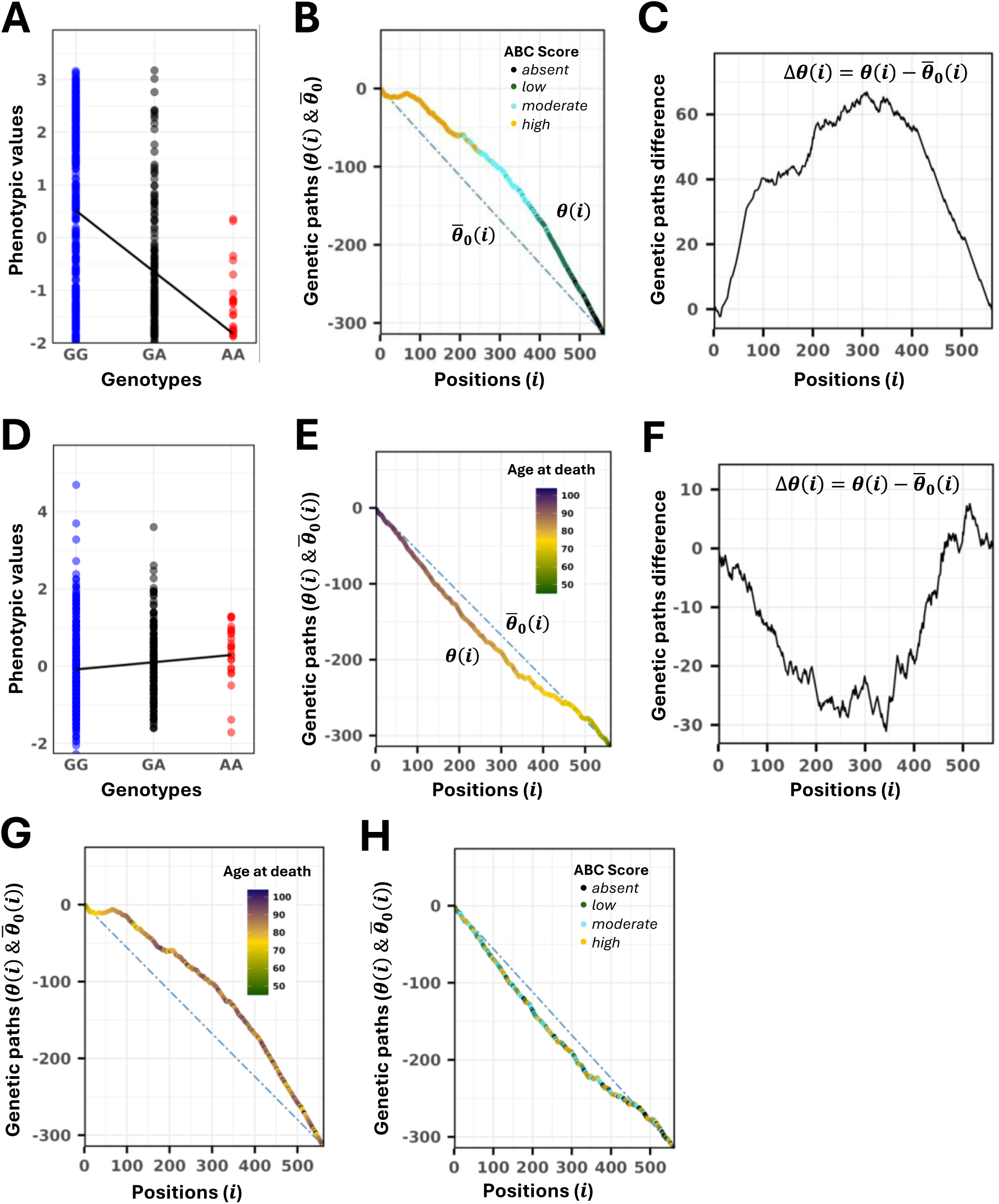
GWAS and GIFT representations of associations: **(A)** GWAS representation of the allelic substitution in rs769449 (Chr19) based on PC1AD+Age. **(B)** GIFT representation of rs769449 using on that PC1AD+Age. Two genetic paths are plotted, one associated with the phenotypic values (θ(i)) and the second not associated with the phenotypic values (θ̄_0_(i)). **(C)** GIFT representation of the difference between the genetic paths determined in (B) used to compute the level of significance underscoring the association. **(D)** GWAS representation of the allelic substitution of rs769449 (Chr19) based on that PC2AD+Age. **(E)** GIFT representation of rs769449 using on that PC2AD+Age. Two genetic paths are plotted, one associated with the phenotypic values ( θ(i) ) and the second not associated with the phenotypic values ( θ̄0(i) ). **(F)** GIFT representation of the difference between the genetic paths determined in (E) used to compute the level of significance underscoring the association. **(G)** Genetic paths from (B) overlaid with age at death measures. **(H)** Genetic paths from (E) overlaid with AD pathology variables.

In Fig 4A, when PC1AD+Age is considered, the allelic substitution effect, based on the partitioning of phenotypic values by genotype as modelled by GWAS, is clearly visible. Fig 4B shows the genetic paths in the GIFT framework, highlighting a clear deviation of θ(i) from θ̄0(i). This deviation is further visible when their difference is considered, as reflected by the large-amplitude paraboloid shape of Δθ(i) in Fig 4C that is used to calculate level of significance of the association. A paraboloid shape with a large amplitude (Fig 4C) arising from ranking phenotypic values, indicates that the different genotypes are not randomly distributed across the whole range of phenotypic values but are segregated in opposite direction within specific subranges, as shown in Fig 4A using the GWAS methodology. Colouration of points with ABC pathology ordinal scales demonstrates the clear correlation of PC1AD+Age with clinical pathology outcomes

When age at death (PC2AD+Age) is considered, the allelic substitution effect, which is negligible or absent in the GWAS framework (Fig 4D), reflects the lack of a clear distinction between the average phenotypic values across genotypes. However, the genotypes are not randomly distributed across the entire range of phenotypic values and show some segregation within specific subranges (e.g., genotype AA in Fig 4D). The genetic paths (Fig 4E) and their differences (Fig 4F) capture this non-random configuration suggesting, in turn, its association with age at death, and shown by the colouration of the datapoints age measures.

Finally, to demonstrate that PC1AD+Age and PC2AD+Age specifically capture AD neuropathology variables and age-at-death variables, respectively, age at death was overlaid onto the genetic path θ(i) from Fig 4B, and AD pathology features were overlaid onto the genetic path θ(i) from Fig 4E. As shown in Fig 4G and Fig 4H, respectively, the features become disordered when projected onto the non-corresponding genetic path.

## Discussion

This study has sought to utilise GIFT for a complex human disease where discreet quantitative variables are not available. Instead, principal component analysis has been implemented to turn a categorical ordinal scale of pathology into near individualised data points for quantitative analysis. Using the resultant principal components GIFT has been directly compared to GWAS to assess its ability to identify subtle genetic effects that could be missed in the traditional analyses.

The three neuropathological features for AD post-mortem confirmation (Thal, CERAD and Braak) alongside age-at death were entered into principal component analysis. The pathology only analysis, yielded PC1AD that captures much of the variance (84.6%); interestingly the additional of age-at-death, decrease the variance captured from the first component (PC1AD+Age) to 64.1%, but still aligned strongly with the pathological features. PC2AD+Age in this analysis captured a further 24.9% and aligned strongly with age-at death, suggesting that age might be less correlated with AD pathology than previous thought. Age is considered a crucial co-variable in dementia analysis given it is the largest risk factor for the disease, doubling risk for AD every 5 years after the age of 65 (18); yet the presence of dementia can shorten life span considerably (19).

The *APOE* locus was associated in all three PC measures analysed, although the significance of these SNPs when associated with pathological traits were greater when age included in the analyses. This result reveals indirectly GIFT’s sensitivity. Indeed, it is notable from Table 2, i.e., using GIFT, that the levels of significance of SNPs can differ between the two PC1-derived traits, PC1AD and PC1AD+Age, as some SNPs are associated specifically with PC1AD, while others are associated with PC1AD+Age. This discrepancy is, to some extent, expected, for two reasons. The first reason is that the PCA analyses informing AD neuropathology were performed on two distinct datasets; consequently, the first principal components, while globally similar, are mathematically different. The second reason relates to the nature of the AD neuropathology dataset, which is informed by scoring systems that provide numerical trait values that are sometimes identical across individuals in the cohort. This imposes a degree of coarseness in GIFT, resulting in some loss of information, which does not happen when the AD pathology and age at death dataset is used that is richer in information. This distinction is illustrated in Table 2, where the column corresponding to PC1AD+Age, contains a greater number of significant SNPs than column corresponding to PC1AD. An exception is rs7826180 on Chr8, whose significance is higher for PC1AD than PC1AD+Age (Table 2). Nevertheless, the level of significance obtained for rs7826180 (pvalue = 2.04 · 10^−6^, Table 2) remains close to the Bonferroni threshold (pvalue = 1.74 · 10^−6^), making it a strongly suggestive SNP.

### Comparison of GWAS and GIFT associated SNPs with AD pathology

Strikingly, the association analyses of PC1AD and PC1AD+Age using either GWAS and GIFT method yielded the same 10 significant SNPs at the genome-wide level on Chr19 all within a 42kb region surrounding the *APOE* gene (*NECTIN2 – TOMM40 – APOE – APOC1*) and included the rs429358 SNPs that determines *APOE epsilon*. To determine whether any of the significant rsIDs common to GWAS and GIFT had previously been associated with Alzheimer’s disease, the GWAS Catalog was interrogated using “Alzheimer” as the trait term to extract relevant bibliographic references (20). The results, reported in the last column of Table 1, demonstrate that all identified SNPs have previously been associated with Alzheimer’s disease.

Interestingly, a recent GWAS combining three large sample datasets together (n=7804) explored genetic association with several Alzheimer’s disease and dementia-related endophenotypes, using either binary or ordinal scales of pathology severity. Pertinent to the current study several genetic associations were found with amyloid-β plaques (defined as none, mild, moderate, severe), NFT Braak staging and CERAD. Analyses with each of these three traits resulted in *APOE* rs429358 being the most significant finding with additional associations for genes around the *APOE* locus (*NECTIN2, TOMM40 and APOC1*) (21), as found in this study with using PC1AD and PC1AD+Age (Table 1 and Table 2).

It is worth noting, however, that, unlike GWAS that associated SNPs on Chr19 exclusively with AD pathology features, GIFT suggests that a subset of these SNPs is also significantly associated with age at death.

### Relevance of associations with AD neuropathology features identified by GIFT

In addition to the *APOE* locus, GIFT identified additional SNPs where GWAS did not.

As done above, to determine whether any of the significant rsIDs determined by GIFT outside Chr19 had previously been associated with Alzheimer’s disease, the GWAS Catalog was re-interrogated using the same trait term as above (20). None of the significant rsIDs from Table 2 were previously reported as associated with AD, which prompted a direct bibliographic search.

Interestingly, rs6734633 (Chr2; *NRXN1*), rs11721706 and rs10866283 (Chr4; *TAPT1-AS1* and *SORBS2*) and rs5758837 (Chr22; *CYB5R3*) did display nominal significance with AD-pathology confirmed diagnosis in the original GWAS of the BDR cohort (6). Furthermore, nominal significance (p<0.05) for SNPs on Chr12, has been observed for rs2270238 *(KLRD1*) (22) and for rs7312352 (*PPM1H*) (23), while the Chr18 SNP rs2298574 (*CDH2*) was also previously identified as nominally significant (23,24).

Additionally, genes identified through their rsID by GIFT in our analysis have been previously implicated in AD and may provide insight into disease mechanisms. *SORBS2* has been linked to cognitive deficits, cortical amyloid-β accumulation, and an increased Aβ42/Aβ40 (25,26), and has also been reported as a genetic modifier of age at onset of AD (27). Our analysis further highlighted *SOX7-AS1*, which was found to be downregulated in circulating exosomes from AD patients compared with healthy controls (28). Given that *SOX7-AS1* likely negatively regulates *SOX7* expression, its downregulation may lead to elevated *SOX7* levels, consistent with enhanced neuronal apoptosis. This aligns with prior studies showing that *SOX7* overexpression promotes apoptosis, whereas *SOX7* knockdown protects neuronal cells from cell death in mouse models (29).

Other notable genes include *PINX1* previously associated with late-onset AD through meta-analysis of GWAS datasets (30), and *MET*, which encodes the hepatocyte growth factor receptor, a tyrosine kinase receptor mediating neurotrophic and neuroprotective signalling (31). Reduced *MET* expression in the cerebral cortex and hippocampus has been observed in AD (32–34).

Immune-related genes were also highlighted. *KLRD1* is a marker of natural killer cells that has been linked to neuroinflammation in AD through its overexpression in human peripheral blood mononuclear cells (35).

Genes involved in neuronal signalling and synaptic function were also identified. *ANO4*, a calcium-dependent non-selective cation channel is also a lipid scramblase expressed in the central nervous system and endocrine tissues associated with cognitive decline in AD (34). *CDH2* encodes N-cadherin, a key cell adhesion molecule for synapse maintenance and neuronal migration, has been associated with amyloid-β–induced synaptic impairment in post-mortem AD brains (36,37). Finally, *CYB5R3* a membrane-bound redox enzyme crucial for cellular energy metabolism and redox balance, has been associated with AD progression (38).

Collectively, these findings demonstrate that gene names identified by GIFT are implicated in multiple pathways relevant to AD pathology.

### Relevance of associations with age at death identified by GIFT

Comparison of the results presented in Tables 1 and 2 shows that GWAS identifies a single SNP from an unnamed gene associated with age-at-death variables (Chr7: rs6952104; PC2AD+Age in Table 1), which is not detected by GIFT. However, this SNP has been linked to genes RNASSP228 and *STK31*, and has previously been identified in GWASs of cognitive performance (39,40).

In contrast, GIFT identifies multiple significant SNPs that are not captured by GWAS. To determine whether any of the significant rsIDs identified by GIFT had previously been associated in relation to a phenotype similar to age at death, the GWAS Catalog was re-interrogated using “life span” as the trait term (20), only returning rs769449 on Chr19 and within the *APOE* region (41). Note that any other trait terms like “health span” or “aging” did not return any result. Interestingly, one of the most significant SNPs for PC2AD+Age, rs13217066 on Chr6, has been linked to the *TBX18* gene involved in cardiac function, including regulation of diastolic blood pressure (42), which is impacted with age (43).

Taken together the association results gained from PC2AD+Age in GIFT highlighted multiple genes connected with various aging processes, supporting the speculation that this component maps onto the age at death variable. Interestingly, while GWAS have shown that *EHBP1* is associated with corpus callosum volume architecture (44), and delayed recall memory performance in AD patients (45), *EHBP1* is also a key regulator of lipophagy, i.e, the depletion of cytosolic lipid droplets, which reverses age-dependent loss of endoplasmic reticulum (ER) proteostasis and enables ER morphological remodelling. This process, in turn, has been shown to drive beneficial metabolic changes and extends the lifespan in C. elegans (46).

Additionally, several genes highlighted affect mechanisms that have also been linked to dementia. *CLCN7* and *ERICHB* are associated with bone density (47) and maculopathy (48), processes linked to aging in the bone and eye respectively, and both low bone density and macular dystrophy have been linked to dementia (49,50). Similarly, genes *EPCIP* and *SGPL1* although have not been linked to age-related diseases, play key roles in processes known to influence aging. The *EPCIP* gene encodes the Exosomal Polycystin1 interacting protein, with is involved it the removal of senescent mitochondria, an organelle implicated in both aging (51,52) and dementia (53), along with exosomes themselves (54). Sphingolipid metabolism has also been linked with the aging process (55); *SGPL1* encodes the enzyme, Sphingosine-1-phosphate lyase 1, which breaks down S1P. A build-up of S1P has been shown to be neurotoxic in mice (47), and associated with cognitive decline (56).

Interestingly other genes highlighted by PC2AD+Age have been directly linked dementia and frailty traits. *MEF2C* was found to be significantly associated with AD in the first GWAS (22); however, this association has been missed in subsequent larger GWAS. Despite this *MEF2C* has been associated with aging, with evidence that its expression decreases with age and with it, its ability to “switch off” microglial to lower inflammation (57). Further to this in a recent GWAS for frailty traits, *MEF2C* was again identified as being associated with poorer cognition (58). In the same GWAS of frailty traits *CDH2* was highlighted as associated as a general factor (58). Neurotrimin encoded by the *NTM* gene is known to play a role in neuronal cell adhesion and has an intronic SNP (rs73035809) associated with tau pathology burden (59).

To conclude, applying GIFT framework to human Alzheimer’s disease–related traits and using principal component–derived traits enabled a more nuanced exploration of genotype–phenotype relationships. Our findings demonstrate three key points. First, GIFT successfully recapitulated the canonical *APOE* locus associations identified by GWAS. This concordance confirms that GIFT retains sensitivity to established, genome-wide significant signals and does not generate spurious associations. Second, GIFT identified additional SNPs beyond those detected by GWAS, including variants in genes with documented links to AD neuropathology. Third, GIFT revealed associations with age at death–related variation that were missed by GWAS. Although methodologically distinct, GIFT and GWAS are rooted in the same scientific tradition. Unlike many machine-learning approaches requiring large amount of data and often characterised as “black boxes” due to limited interpretability, both GIFT and GWAS are transparent analytical frameworks in which assumptions and inferential procedures are explicitly defined. In this sense, GIFT constitutes a complementary and interpretable methodology capable of enhancing inference in contexts where large sample sizes are impractical but phenotypic resolution is high, thereby helping to mitigate longstanding power limitations in quantitative genomics.

## Materials and methods

### Dataset

The genotyping and quality control on the BDR cohort is detailed elsewhere (7). This dataset comprised 563 BDR participants with complete neuropathological assessments for Alzheimer’s disease (AD), including CERAD score, Thal phase, and Braak tau stage, which together form the ABC score for AD neuropathological change. The Consortium to Establish a Registry for Alzheimer’s Disease (CERAD) score is a semi-quantitative amyloid rating scale (0–3) used to classify the density of neuritic β-amyloid plaques in the brain. Thal amyloid staging (0–5) characterises the hierarchical anatomical spread of β-amyloid deposition. Stage 0 indicates no amyloid deposits. Stage 1 reflects deposits confined to the neocortex, stage 2 indicates spread to allocortical regions (e.g., hippocampus), stage 3 reflects involvement of the thalamus and striatum, stage 4 indicates extension to the brainstem, and stage 5 denotes amyloid deposition in the cerebellum in addition to all previously affected regions. Braak tau staging (0–VI) categorises disease progression based on the distribution of neurofibrillary tangles (NFTs). Stage 0 indicates no NFTs. Stage I involves the medial temporal lobe. Stage II reflects spread to the entorhinal cortex and hippocampal CA1 region. Stage III involves the hippocampus, amygdala, and early neocortical regions. Stage IV indicates further spread to temporal, parietal, and occipital cortices. Stage V reflects widespread involvement of association neocortical areas, and Stage VI denotes NFTs in primary motor and sensory cortices, indicating extensive cortical involvement. Stages I–III are typically associated with preclinical AD (no significant clinical symptoms), Stages III–IV with prodromal disease often diagnosed as mild cognitive impairment, and Stages V–VI with clinically manifest dementia. These three neuropathological measures (CERAD, Thal phase, and Braak stage) are combined into the ABC score to determine the overall level of Alzheimer’s disease neuropathological change. ABC classifications of “Intermediate” or “High” are considered sufficient to account for the observed dementia (8).

### Generation of Quantitative Phenotypes

The ABC score for AD and, the ABC score for AD with or without age at death, were entered into a principal component analysis (PCA) to produce to set of quantitative independent variables for analysis using samples with sufficient genotype information (missingness ≤ 10% ). Principal components with or without age at death were entered into traditional GWAS quantitative trait association analyses and GIFT to detect SNPs related to AD.

### Association Analysis

Typical GWAS methodologies were employed alongside GIFT to perform association analysis. Genetic data were filtered to include individuals of interest in the study; we removed samples with no corresponding phenotypic information for Thal, CERAD and Braak Tangle as well as for missing pathology group information. The genetic data were also filtered for biallelic SNPs, selecting minor allele frequency (*MAF* ≥ 5%), missingness per SNP ≤ 10% and HWE ≤ 10^−6^. Genetic data were aligned to the hg19 reference genome, stored at UCSC genome browser database (9), and rsIDs were imported from the latest dbSNP data for the same hg19 reference (10). This filtering and formatting was accomplished using bcftools, vcftools and plink software (www.cog-genomics.org/plink/2.0/) (11–13). GWAS pvalue calculations were performed using GEMMA software (14). Kinship matrices were not included in the GWAS and GIFT analyses for it was previously shown that the BDR cohort is from European ancestry (15).

### GIFT

The filtered genetic data were passed into the GIFT algorithm where genotypes were first transformed as follow, (0|0) →(+1), (0|1 and 1|0)→(0) and (1|1)→(-1), and subsequently positioned alongside ordered phenotypic values (smallest to largest), then the cumulative sum of genotypes were computed and represented under the form of a genetic path whose amplitude was used to compute the level of significance between the genotype and the phenotype as detailed elsewhere (4). The GIFT methodology will be introduced in the main text.

### Significance threshold for GWAS and GIFT

To calculate Bonferroni thresholds for the GWAS and GIFT results, plink 2.0 (11) was used to clump the association data using the filtered genotype data as a reference to keep only the most significant and non-LD SNPs (see S1 Tables). The filters used were clump-r2 of 0.1, clump-kb of 250, and clump- p1 of 0.05. The total number of index SNPs were used to calculate 5% Bonferroni threshold as standard (-log10(0.05/Nind-SNPs) per analyses.

### Computation of significance for GIFT

As detailed elsewhere (4), for each SNP the maximal amplitudes resulting from the cumulative sum of genotypes ranked as a function of phenotypic values were used to compute the level of significance of associations.

### Software and codes

All figures were designed in R software, using ggplot package. All scripts are run on the University of Nottinghams Ada HPC cluster service utilising the slurm environment and conda environments created under conda version 23.7.4, bioconda and conda forge (16,17). The code for GIFTv2.0 is given as supporting information S2.

## Data Availability

All data produced in the present work are contained in the manuscript

https://www.brainsfordementiaresearch.org/

## Data and code availability

Source data for this study are publicly available either from the UK Brain Banks Network (https://ukbbn.brainsfordementiaresearch.org/public/en/) or the Dementias Platform UK (https://www.dementiasplatform.uk/). Supporting information S1 Tables and S2 Software are available on https://doi.org/10.6084/m9.figshare.32063487 and https://doi.org/10.6084/m9.figshare.32063703, respectively.

## Acknowledgements

We would like to gratefully acknowledge all donors and their families for the tissue provided for this study. Human post-mortem tissue was obtained from the South West Dementia Brain Bank, London Neurodegenerative Diseases Brain Bank, Manchester Brain Bank, Newcastle Brain Tissue Resource and Oxford Brain Bank, members of the Brains for Dementia Research (BDR) Network. The BDR is jointly funded by Alzheimer’s Research UK and the Alzheimer’s Society in association with the Medical Research Council. We also wish to acknowledge the neuropathologists at each centre and BDR Brain Bank staff for the collection and classification of the samples. Brains for Dementia Research has ethics approval from London – City and East NRES committee 08/H0704/128+5 and has deemed all approved requests for tissue to have been approved by the committee.

