## Supplementary material for "Identifying single-nucleotide polymorphisms (SNPs) intersecting Alzheimer’s disease pathology and end-of-life traits using genomic informational field theory (GIFT)": GIFTv.2 code: Workflow_AD.html (from another version of this preprint: version 2, 2026-03-30)

### GIFT vs GWAS subsampling and testing

This file gives a summary of the workflow of analysis, describing the scripts run in order and giving example settings to get certain outputs.
All work was completed using the University of Nottingham ADA HPC systems. Scripts were re-used for both PC1-4 data and PC1-3 data, using different settings for each dataset and each output within them. An example of settings used are shown, but scripts can be run with different settings to get different outputs.

---

#### Key:

Figure contribution: 1A-Z = The script helps create the specified figure in final writeup (A-Z is placeholder as subfigure labels can vary within this range)

---

### PREP STAGE

#### setup

- First we create a new project directory where all scripts and data will be stored
  - in my case this is : "/gpfs01/home/brash79/PROJECTS/AD\_UNIMPUTED\_PC1-4\_Control\_plus\_AD" for working on PC1-4 data where we included the "Age of death" as a component, or "/gpfs01/home/brash79/PROJECTS/AD\_UNIMPUTED\_PC1-3\_Control\_plus\_AD" where "Age of death" was not considered in the PCA.
- Inside the project directory we create the following folders:
  batch\_files, core\_files, output\_files, prep
- Save all scripts required (see starting files below) to the "batch\_files" folder
- Save all data files inside of "prep" folder

---

#### starting files

Data files:

- Phenotype and genotype data sourced through Brains for Dementia Research
- Phenotype file 1: Phenotype\_Info.csv
- Phenotype file 2: BDR\_Sample\_Demographics.csv
- genotype file 1: GIFTsamples (.bim .fam. bed)

Other files sourced online ( with their formats modified locally where necessary):

- HG19 FASTA Reference file : hg19.nochr.fa
  - removed the "chr" before each chromosome number -> (1,2,3...22)
- DBSNP Genotype file : dbsnp\_hg19\_ready\_vcf.gz

- reformatted chromosome naming scheme to just numbers as well (1,2,3...22)

- HG19 Known gene GTF

Script files:

- script 1: download\_files\_ADA.sh
- script 2: Make\_Phenotype\_AD\_05\_12\_25.R
- script 3: filter\_vcf\_ADA.sh
- script 4: stage\_1\_ADA.sh
- script 5: stage\_1.py
- script 6: run\_and\_monitor\_ADA.sh
- script 7: run\_and\_monitor.py
- script 8: GIFT\_CORE.py
- script 9: GIFT\_V1.0.py
- script 10: convert\_GWAS\_format.py
- script 11: stage\_2\_ADA.sh
- script 12: stage\_2\_1.py
- script 13: make\_plink\_LD\_script.R
- script 14: calc\_thresholds.py
- script 15: stage\_2\_2.py
- script 16: stage\_2\_MANHATTAN\_ADA.sh
- script 17: stage\_2\_MANHATTAN.py
- script 18: Rscript\_run\_and\_monitor.py
- script 19: stage\_3\_ADA.sh
- script 20: stage\_3\_crossreferencing.py
- script 21: U\_biomart\_translate.R
- script 22: U\_stringDB.R
- script 23: stage\_3\_UPSET.R
- script 24: stage\_5\_ADA.sh
- script 25: stage\_5\_fetch\_data.py
- script 26: stage\_5\_ram.py
- script 27: U\_plot\_genotype.R

- Scripts not listed here were not used in the creation of figures or data tables for the paper, OR were created from one of the scripts inside the list and ran based on that scripts input values.


---

#### 1 Install software/files and setup environment

- Firstly, we create the conda environment "gift\_env" required for analysis by including the packages listed inside conda\_environment\_setup.sh
- Other software is provided by ADA through modules, these modules are:
  - module load anaconda-uoneasy/2023.09-0 (conda)
  - module load plink-uoneasy/2.00a3.7-foss-2023a (plink 2)
  - module load plink-uoneasy/1.9b\_6.21-x86\_64 (plink 1.9)
  - module load vcftools-uoneasy/0.1.16-GCC-12.3.0 (vcftools)
  - module load bcftools-uoneasy/1.18-GCC-13.2.0 (bcftools)
- GEMMA was installed separately into a folder called software\_installs
  - Version: = 0.98.5 (2021-08-25)
- HG19 fasta reference file, DBSNP file and HG19 knowngene GTF were downloaded using download\_files\_ADA.sh

#### 2 Quick filter of Genotype file

- Inside the "prep" folder I ran the command "plink2 --bfile GIFTsamples --mind 0.10 --make-bed --out GIFTsamples\_mind\_filter"
  - This gave genotype bed files where samples had plenty of genotype information to work with.
  - The IDS inside the .fam file would be passed to the R script "Make\_Phenotype\_AD\_05\_12\_25.R"

#### 3 Creating the phenotypes to work with

- Next we run "Make\_Phenotype\_AD\_05\_12\_25.R" with the following variable settings
- for PC1-4 data:
  - PCs\_to\_analyse <- 4
  - Include\_controls <- yes
- for PC1-3 data (Age of death excluded):
  - PCs\_to\_analyse <- 3
  - Include\_controls <- yes
- This script will output:
  - the phenotype files "\*\_phenotype.csv"
  - the phenotype labelling data
  - the Keep file of individuals we want from the genotype files
  - the screeplots: **Figure contribution (2A-B)**
  - the biplots : **Figure contirbution (2C-D)**

#### 4 Filtering genotype information

- Now I dealt with the genotype data once I knew which samples I would need, applying the following settings inside the script "filter\_vcf\_ADA.sh" :
  - PTU="/gpfs01/home/brash79"
  - downloads="${PTU}/downloads"
  - start\_files="AD\_geno\_prepared"
  - start\_file\_type="vcf"
  - reference\_version="HG19"
  - required\_version="HG19"
  - need\_rsID="yes"
  - fixreference="no"
  - MAX\_ALLELES=2
  - MIN\_ALLELES=2
  - MAF=0.05
  - MISS=0.9
  - HWE\_VAL=1e-6
  - filter\_samples="no"
  - samples\_filter\_file="samples\_to\_keep.txt"
- This leaves me with 242105 SNPs and 598 samples (563 needed for analysis only)

#### 5 Setting up the analysis runs

- Using stage\_1\_ADA.sh we can set up the scripts to run with the following settings
  - PTM="/gpfs01/home/brash79/PROJECTS/AD\_UNIMPUTED\_PC1-4\_Control\_plus\_AD"
  - vcf\_file="${PTM}/core\_files/FINAL\_AD\_geno\_prepared.vcf"
  - PTU="/gpfs01/home/brash79"
  - environment="gift\_env"
  - total\_snps\_file="${vcf\_file%.vcf}\_total\_snps.txt"
  - custom\_id\_column="Accession\_ID"
  - GiftOrGwas="BOTH"
  - server="ADA"
  - cores=6
  - phenotype\_names="PC1 PC2 PC3 PC4"
- This will create bash scripts for each phenotype file where GIFT and GWAS will run on that phenotype and genotype data (genotype data is shared amongst runs; samples are the same across runs)

---

### ANALYSIS STAGE

#### 6 Running GIFT/GWAS calculations

- run\_and\_monitor\_ADA.sh will run all scripts in the project file "parallel" folder using run\_and\_monitor.py with the following settings for example:
  - PTU="/gpfs01/home/brash79"
  - job\_count=10 (maximum number of parallel jobs)
  - PTM="/gpfs01/home/brash79/PROJECTS/AD\_UNIMPUTED\_PC1-4\_Control\_plus\_AD" (for PC1-4 data)
- Analysis will follow through the bash scripts (as written by stage\_1.py) using the following scripts in this order

1. GIFT\_CORE.py -> to transform genotypes into GIFT readable format (-1,0,1,NA) and to orders them by phenotype
2. GIFT\_V1.0.py -> to run GIFT on the outputs of GIFT\_CORE.py using compressed numpy genotype matrix
3. /gpfs01/home/brash79/software\_installs/gemma ->calculate GWAS values
4. convert\_GWAS\_format.py -> convert Gemma GWAS output to match GIFT output for collecting data in the following step.

- Now we have the following results:
  - output\_files/GWAS\_RESULTS/ -> all GWAS results for each phenotype
  - output\_files/GIFT\_RESULTS/ -> all GIFT results for each phenotype
  - output\_files/permutation\_results/ -> GIFT permutation runs for calculating permutation threshold (if needed)

#### 7 Collecting GIFT/GWAS results and creating SNP\_DATA files

- To obtain these files we first must run stage\_2\_ADA.sh with the following settings for example:
  - PTM="/gpfs01/home/brash79/PROJECTS/AD\_UNIMPUTED\_PC1-4\_Control\_plus\_AD"
  - vcf\_file=${PTM}/core\_files/FINAL\_AD\_geno\_prepared.vcf
  - reference\_prefix="FINAL\_AD\_geno\_prepared"
  - SNP\_name='rsID'
  - PTU="/gpfs01/home/brash79"
  - server="ADA"
  - env="gift\_env"
  - jobListDf="${PTM}/core\_files/JOB\_LIST.csv"
- Plink2 snp clump settings (also set inside of stage\_2\_ADA.sh)
  - threshold\_value=0.05
  - r\_square=0.1
  - range\_kb=250
- This bash script will use the following scripts in this order:

1. stage\_2\_1.py -> gather the data and attach information like rsID, ref alt etc
2. make\_plink\_LD\_script.R + stage\_plink\_filter\_LD\_ADA.sh -> create and run plink clumping scripts on the snps
3. calc\_thresholds.py -> calculate thresholds e.g. bonferroni 5%, 1% etc...
4. stage\_2\_2.py -> Join the clump data to SNP\_DATA files and fetch number of genostates (-1,0,1, NA) -> (Nm,Nz,Np,Nmiss).

- We now have tsv files on SNP results for GIFT and GWAS for EACH phenotype we ran the samples against. These are stored in output\_files/SNP\_DATA

#### 8 Manhattan plotting

- To generate manhattan plots we run stage\_2\_MANHATTAN\_ADA.sh with the following settings for example:
  - vcf\_file="FINAL\_AD\_geno\_prepared.vcf"
  - server='ADA'
  - sourceFile="SNP\_DATA" (change this to "LD" when looking at LD snps only)
  - threshold="bf\_0.05" (change this to bf\_0.01 to look at 1% bonferroni)
- This will run scripts in the following order:

1. stage\_2\_MANHATTAN.py -> Write the R scripts in python based on given settings for GWAS and GIFT data
2. Rscript\_run\_and\_monitor.py -> Runs the bash scripts in parallel

- Inside output\_files/stage\_2\_output/ we now have our GIFT and GWAS graphs **Figure contribution (3A-C)**

#### 9 Gene analysis

- To generate gene lists, we may run stage\_3\_ADA.sh with the following settings for example:
  - PTM="/gpfs01/home/brash79/PROJECTS/AD\_UNIMPUTED\_PC1-4\_Control\_plus\_AD"
  - PTU="/gpfs01/home/brash79"
  - genome\_ref\_file="hg19.knownGene.gtf"
  - server="ADA"
  - environment="gift\_env"
  - crossreference\_output="${PTM}/output\_files/crossreference\_output"
  - sort\_genes\_input="${PTM}/core\_files/sort\_genes\_input"
  - sort\_genes\_output="${PTM}/output\_files/sorted\_genes"
  - biomart\_input="${PTM}/core\_files/biomart\_input"
  - biomart\_output="${PTM}/output\_files/biomart\_output"
  - stringDB\_input="${PTM}/core\_files/stringDB\_input"
  - stringDB\_output="${PTM}/output\_files/stringDB\_output"
  - upset\_input="${PTM}/core\_files/upset\_input"
  - sort\_genes\_output\_venn\_diagrams="${PTM}/output\_files/sort\_genes\_venn\_diagrams"
  - stage\_3\_output\_folder="${PTM}/output\_files/stage\_3\_output"
  - biomart\_mode="BOTH" # OPTIONS [GIFT | GWAS | BOTH]
  - gene\_limit=2000
  - data\_type="uncorrected"
  - thresh\_name=bf\_0.05
- stage 3 will run scripts in the following order:

1. stage\_3\_crossreferencing -> select snps to crossreference with genes and make a bedtools bash script
2. cross\_reference\_script.sh -> uses bedtools to intersect between chosen GTF and snp list
3. U\_biomart\_translate.R -> translates IDs into gene names if available
4. U\_stringDB.R -> runs string analysis on gene lists (not required for our paper)
5. stage\_3\_UPSET.R -> organises genes into phenotype categories, showing any overlaps between phenotypes (not required for our paper)
6. U\_stringDB.R -> re-ran on those sorted categories (not required for our paper)

- This set of inputs will give us genes from snps that are above the 5% bonferroni threshold based on their GIFT or GWAS pvalue (considering each method separately). These are stored inside output\_files/stage\_3\_output/ .
- **Figure contribution (TABLE 1, TABLE 2)**

#### 10 Pathway analysis

- To generate the genetic path plots from GIFT we run stage\_5\_ADA.sh with the following settings for example:
  - PTU="/gpfs01/home/brash79"
  - server="ADA"
  - jobListDf="${PTM}/core\_files/JOB\_LIST.csv"
  - s5Data="${PTM}/core\_files/s5\_Data"
  - s5Out="${PTM}/output\_files/stage\_5\_output"
  - batch\_startFolder="${PTM}/batch\_files/s5\_batch\_start"
  - PTM="/gpfs01/home/brash79/PROJECTS/AD\_UNIMPUTED\_PC1-4\_Control\_plus\_AD"
  - VCF="FINAL\_AD\_geno\_prepared.vcf"
  - pheno\_label="YES"
  - dtype="uncorrected"
  - rsID\_names="rs6952104"
- stage\_5\_ADA.sh will run scripts in the following order:

1. stage\_5\_fetch\_data.py -> selects the snps to get paths of and makes bash scripts to analyse their genetic paths
2. stage\_5\_ram.py -> runs the bash scripts created from previvous python script

- Simply replacing the rsID\_names variable for any rsID available in the dataset will allow you to investigate that snps genetic path.
- These are stored inside output\_files/stage\_5\_output/ .
- **Figure contribution (4A-H)**

---
